# Proposed Classification System for the 445 nm Blue Light Laser for Treatment of Laryngeal Lesions

**DOI:** 10.64898/2026.04.20.26351290

**Authors:** Mahnoor Khan, Atif M. Islam, Yassmeen Abdel-Aty, David Rosow, Pavan Mallur, Michael Johns, Clark A. Rosen, Yael E. Bensoussan

## Abstract

**Objective:** Only preliminary investigations on the use of the 445 nanometer wavelength blue light laser (BLL) for various laryngeal pathologies have been described. Currently, no standard exists for reporting treatment technique and tissue effect with this modality. Here, we aim to establish and validate a classification system to describe laser-induced tissue effects.

**Study Design:** Retrospective video-based study for classification development and reliability validation.

**Methods:** Video recordings from procedures performed with the BLL by multiple academic laryngologists were retrospectively reviewed. A preliminary 6-point classification (BLL 1-6) was developed based on expert consensus. Thirteen additional procedural clips were independently rated utilizing the classification schema to assess perceived tissue effect, and measure inter- and intra-rate reliability.

**Results:** The final 5-point classification system (BLL 1-5) included angiolysis, blanching, tissue vaporization, ablation with mechanical tissue removal, and cutting. The consensus of the combined reviewers in rating all cases was 89% (58 of 65). Complete consensus was not achieved in 11% (7/65) of cases. Of those incorrect, 57% (4/7) were of clips illustrating the BLL-2 classification. Intra-rater reliability amongst the reviewers was 100%.

**Conclusion:** Tissue effect of the 445 nm blue light laser can reliably be standardized with this proposed classification system. This rating system can be used to facilitate future systematic study of outcomes and effective communication between laryngologists and trainees.

## INTRODUCTION

Otolaryngologists were amongst the first specialties to adapt lasers after their introduction to the medical community in the 1960s.^1^ The CO_2_ laser was the initial model of choice after the it was found to be favorably absorbed by almost all human body fluids and soft tissue due to its 10,600 nanometer (nm) wavelength, the wavelength at which water is absorbed.^1^ Bredemeier’s endoscopic delivery system, developed in 1967, allowed for in vivo canine studies of the laser by Jako.^2,3^ Following these landmark developments over 50 years ago, the era of modern laryngeal laser surgery was ushered in.

While CO_2_ remained a mainstay because of its efficient cutting properties, other lasers were developed with varying abilities. Photoangiolytic laser surgery was developed, encompassing the pulsed dye laser (PDL), at a 585 nm wavelength, and the potassium titanyl phosphate (KTP) laser, at a 532 nm wavelength. Both lasers allowed for photocoagulation due to their predominant absorption by oxyhemoglobin, thus, they proved ideal for vascular lesions i.e. vocal fold ectasias, varices, and hemorrhagic lesions.^4^ Additionally, their flexibility and relative portability allowed for cost effectiveness. The KTP laser was temporarily discontinued in 2018, creating a gap in the market for alternative laser technologies.

In 2020, the novel 445 nm blue light laser (BLL) was introduced in the United States. Initial efficacy studies of the laser showed that it was comparable to CO_2_ lasers and KTP lasers.^5^ Similar to the KTP laser, the 445 nm blue light laser provides photoangiolytic capabilities, with oxyhemoglobin serving as the primary tissue chromophore, enabling vessel ablation and coagulation at low energy levels, with tissue ablation and carbonization at higher energy levels. It proved effective at treating vascular ectasia, papilloma, Reinke’s edema, polyps, contact granuloma, and other benign lesions in the office and during general anesthesia.^6^ Unlike the KTP, the blue light laser also provides cutting capability, with thin coagulation margins that minimize collateral tissue injury.^5^ Animal studies looking at rat larynges treated with KTP and blue light laser reported significantly decreased protein deposition and fibrosis when compared at the 3 month mark, suggesting reduced scarring with the blue light laser.^7^ Because of the similar clinical utility and concern for KTP availability in the future, laryngologists have increasingly turned to the blue light laser.

A validated classification system for the tissue effects of the KTP laser has been developed and validated.^8^ However, no similar standardized classification system exists to report the tissue effects of the blue light laser.

The aim of this study is to establish a new classification system to describe the desired tissue effect using the 445 nm blue light laser for laryngeal surgery and to validate the classification schema through inter- and intra-rater reliability metrics.

## METHODS AND MATERIALS

Video clips of 445 nm blue light laser procedures from multiple institutions were retrospectively reviewed by six fellowship-trained academic laryngologists. Ethical approval to report this study was obtained from our university’s Institutional Review Board (USF IRB STUDY004249), and the requirement to obtain informed consent was waived as the study was considered a review of existing data.

### Retrospective Review of Videos of 445 nm Blue Light Laser Treatment

Video clips of blue light laser laryngeal treatments were retrospectively reviewed from four separate academic institutions; University of South Florida, University of Miami, Beth Israel Deaconess Medical Center, and the University of Southern California. Video clips included pathologies such as vocal fold nodules, cysts, polyps, recurrent respiratory papillomatosis, webbing, ectatic vessels, and subglottic stenosis. All clips were from treatments performed either awake with a working channel distal tip laryngoscope, or in the operating room through micro suspension laryngoscopy under general anesthesia.

### Creation of the Classification System Through Expert Consensus

A novel classification system describing the treatment effect from the BLL was developed through expert consensus by six academic laryngologists based on iterative review of the video clips described above. After 5 hours of expert consensus over 3 sessions, it was agreed that the outcome desired was to develop a classification that would help laryngologists describe treatment effect in a standardized manner to improve communication between experts, optimize documentation, and enhance trainee education.

The influence of specific laser settings was considered, however, review of the cases revealed that different pathologies responded variably to these settings. Moreover, since the laser is handheld, the distance between the tip of the laser and the tissue plays an important role in determining the tissue effect. For these reasons, the group concluded that this first classification would only include the tissue effect. Further investigation would be required to establish the laser parameters necessary to reproducibly achieve these tissue effects across varying pathologies.

While reviewing the videos and through discussion, the group also noticed that some cases involved contact of the laser fiber with the tissues while other techniques did not. For that reason, the attribute “contact” (C) or “non-contact” (NC) mode was added to the classification.

### Selection of 445-nm Blue Light Laser Clips to Assess for Intra- and Inter-rater Reliability

Video recordings of patients undergoing treatment with the 445 nm blue light laser were retrospectively selected to include a wide range of tissue effects. Each video clip was edited to show pre-treatment tissue, a portion of the laser treatment, and the post-treatment end-tissue effect. Footage of the laser treatment was included to enable delineation between contact and non-contact technique.

Thirteen video recordings were compiled and presented to reviewers in a single-blind fashion. Reviewers were blinded to the treatment classification, distribution of clips across classifications, and the number of repeated clips. Prior to review, the rating system was provided to each reviewer, along with one identified example demonstrating each BLL classification.

Reviewers were asked to classify each of the 13 clips utilizing the provided classification system of end-tissue effects and contact / non-contact modality. At least one clip of each classification, BLL-1 through 6, was included. Three were selected at random to be repeated; one recording for BLL-6 and two for BLL-2. Reviewers were asked not to review previous clips after selecting the classification to avoid intra-rater bias.

### Data Management and Statistical Analysis

Reviewers accessed the same video clips through a Google Drive file and entered responses into a Microsoft Excel sheet. The primary outcome was inter-rater reliability among laryngologists when classifying video recordings according to the proposed system. Intra-rater reliability was assessed by classification comparison of the three repeated recordings.

## RESULTS

### Preliminary Grading System

Based on expert consensus, a novel 6-point classification system was designed (Table 1). BLL-1, designated angiolysis, is aimed at selective vessel ablation; in this setting, vascular structures are ablated with minimal to no blanching of the surrounding mucosa in either contact or non-contact mode (Fig 1A). BLL-2, designated blanching, results in mucosal blanching without epithelial disruption in both contact and noncontact settings (Fig 1B). BLL-3, designated epithelial disruption, results in epithelial disruption without deeper tissue ablation. BLL-4, designated tissue vaporization, results in tissue ablation deeper than the epithelium in both settings (Fig 1C). BLL-5, designated ablation with mechanical tissue removal, results in ablation and debridement with the tip of the laser exclusively in contact mode (Fig 1D). Finally, BLL-6, defined as cutting, involves linear vaporization to achieve tissue separation in contact and non-contact settings (Fig 1E).

**Table 1.** Preliminary 6-point Classification System for 445 nm Blue Light Laser Tissue Effects.

| **Classification** | **Definition** | **Contact (C) or Non-Contact (NC)** |
| --- | --- | --- |
| **BLL-1: Angiolysis** | Selective vessel ablation | C or NC |
| **BLL-2: Blanching** | Blanching of the epithelium without breach | C or NC |
| **BLL-3: Epithelial Disruption** | Disruption of the epithelium without deeper lesion ablation | C or NC |
| **BLL-4: Tissue Vaporization** | Tissue ablation deeper than the epithelium | NC only |
| **BLL-5: Ablation with Mechanical Tissue Removal** | Mechanical ablation and debridement | C only |
| **BLL-6: Cutting** | Linear vaporization to achieve tissue separation | C or NC |

**Figure 1.**
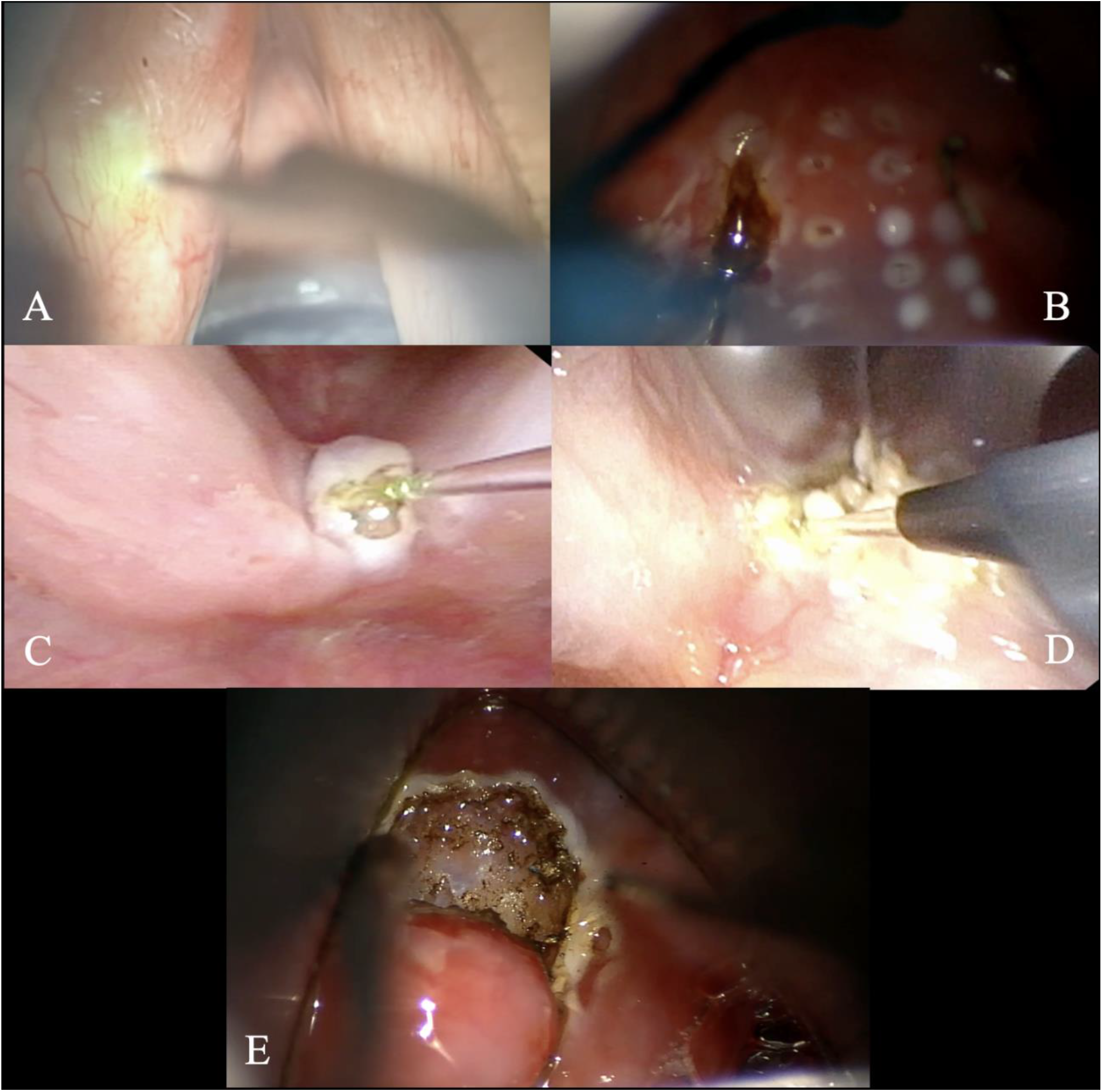
**Figure 1A.**Visual example of angiolysis (BLL-1) **Figure 1B**. Visual example of blanching (BLL-2) **Figure 1C**. Visual example of tissue vaporization (BLL-3) **Figure 1D**. Visual example of ablation with mechanical tissue removal (BLL-4) **Figure 1E**. Visual example of cutting (BLL-5)

### Video-Perceptual Analysis for Inter- and Intra-rater Reliability

Inter-rater consensus across all cases was 89% (58/65), excluding repeat clips. Consensus rates by classification were as follows: BLL-1, 100% (5/5); BLL-2, 80% (16/20); BLL-3, no cases identified; BLL-4, 90% (9/10); BLL-5, 93% (14/15); BLL-6, 93% (14/15) (Figure 2).

**Figure 2.**
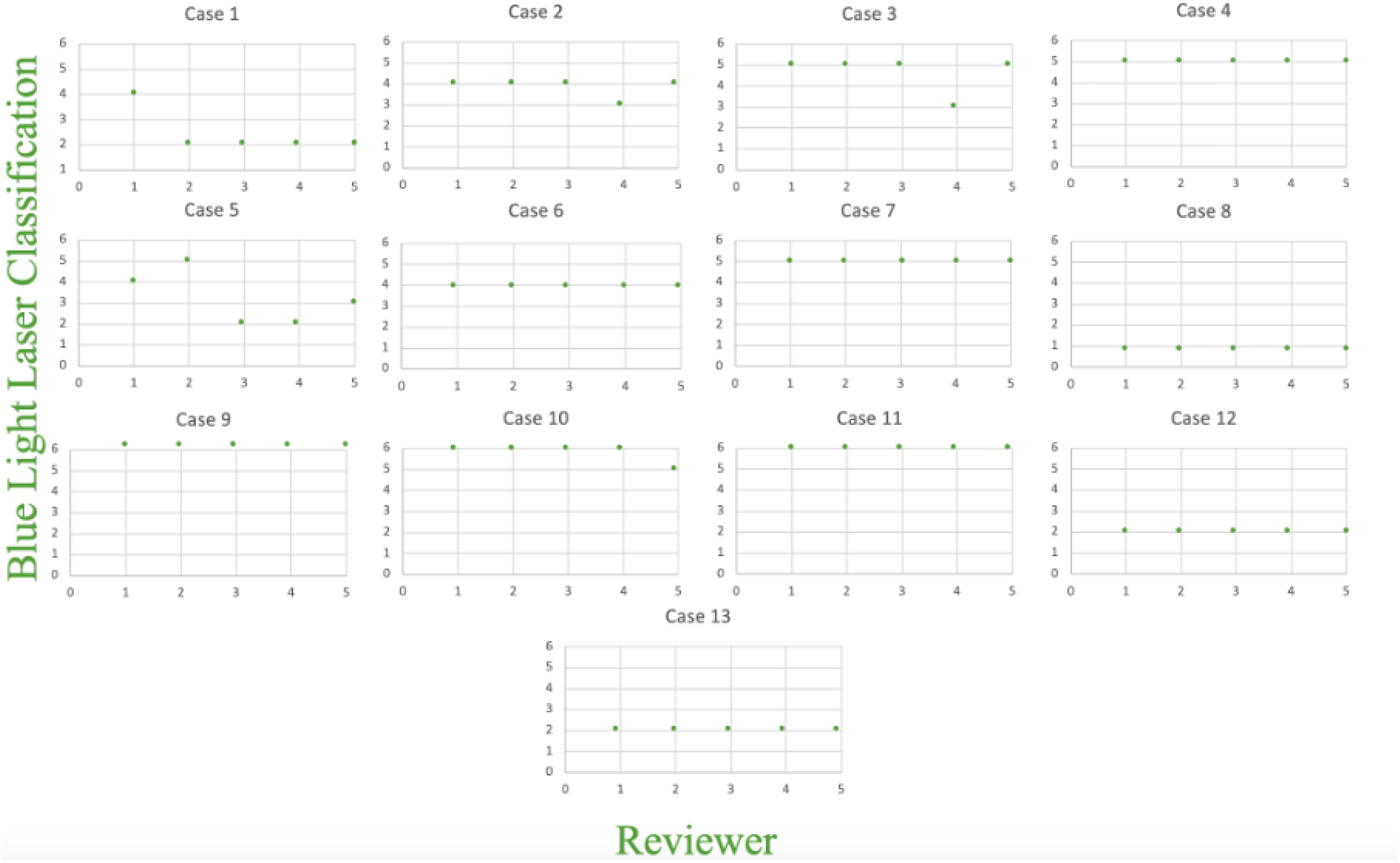
Results of the video perceptual analysis

Complete consensus was not achieved in 11% (7/65) of cases. Of those incorrect, 57% (4/7) were of clips illustrating BLL-2 classification (epithelial blanching). Intra-rater reliability amongst the reviewers was 100%.

### Edited Final Classification Schema

After in-depth review of video clips, the 6-point classification system was challenged. No videos appeared to resolutely fall into the category of BLL-3 (epithelial disruption). A decision was made to remove this class all together. Therefore, our final proposed schema includes only the remaining 5 classes (Table 2).

**Table 2.** Final 5-point Classification System for 445 nm Blue Light Laser Tissue Effects.

| **Classification** | **Definition** | **Contact (C) or Non-Contact (NC)** |
| --- | --- | --- |
| **BLL-1: Angiolysis** | Selective vessel ablation | C or NC |
| **BLL-2: Blanching** | Blanching of the epithelium without breach | C or NC |
| **BLL-3: Tissue Vaporization** | Tissue ablation deeper than the epithelium | NC only |
| **BLL-4: Ablation with Mechanical Tissue Removal** | Mechanical ablation and debridement | C only |
| **BLL-5: Cutting** | Linear vaporization to achieve tissue separation | C or NC |

## DISCUSSION

This single blind video-perceptual analysis demonstrates that independent reviewers can adopt and apply the proposed classification system in a valid manner. Altogether, there was a high consensus rate of 89% and with an intra-rater reliability of 100%. When compared to a similar study by Mallur et al. examining the tissue effects of the KTP laser where inter-rater reliability was determined to be 83%, this inter-rater reliability is comparable.^8^

Consensus in the rating system was lowest when classifying BLL-2 (80% or 16/20). Of the 4 mis-ratings, two were rated as BLL-4, one as BLL-5, and the other as BLL-3 (Figure 2). It is possible that in the contact form of BLL-2, tissue may inadvertently stick to the laser tip thereby making it appear as mechanical tissue removal, a quality seen in BLL-4. This tendency to adhere therefore introduces a minimal limitation of the classification system. In the two remaining misclassifications (cases 2 & 3), clips were designated BLL-3 despite overall consensus ratings of BLL-4 and BLL-5, respectively. After discussion with the reviewers, BLL-3 was agreed to be a transitional effect which may occur with, or slightly before, other effects and rarely as an end-tissue effect. As a result of this determination, the decision was made to redact the original BLL-3, described as epithelial disruption without deeper tissue ablation, thus resulting in a final classification with 5 total categories.

It is important to note that no other classifications systems for the blue light laser currently exist for direct comparison in otolaryngology or any other medical specialty. Furthermore, there is very little comparative data and/or attempts at standardizing the effects of the BLL. Outside of otolaryngology, the BLL has demonstrated applications in dermatology, specifically for the treatment of rosacea and acne.^9^ Additionally, a comparative study completed within the specialty of dentistry examined the effects of BLL versus infrared diode at the cellular level, concluding that the BLL showed improved absorption without major side effects in adjacent tissue.^10^ Therefore, this classification schema may have potential utility in other medical fields beyond otolaryngology.

The importance of this classification system lies in the ability to cohesively communicate and document tissue effects between laryngologists and trainees in both the general anesthesia and awake setting. A crucial byproduct of the classification system is its ability to facilitate future comparative outcomes studies. With the heavy reliance on the blue light laser as of late due to the temporary removal of the KTP laser from the market, it is critical that an established validated method exists to classify the effect of the blue light laser.

### Limitations and Future Directions

Although this study describes tissue effects by the BLL, it does not include proposed laser settings or techniques to obtain that tissue effect. This was discussed in depth during the expert consensus sessions and our group concluded that the type of tissue, type of pathology, and distance from the tissue clearly impacts the laser settings needed to obtain the desired tissue effect. Further prospective work will be needed to correlate the laser settings required to obtain the desired tissue effect for varying laryngeal pathologies. Care should be taken to separate settings in the awake versus general anesthesia setting as the distance from the tissue and the dispersion of the heat from the laser can vary in these different conditions.

## CONCLUSIONS

The proposed classification schema for reporting 445 nm blue laser treatment tissue effects is an effective grading system developed through expert consensus. This single blind video-perceptual analysis demonstrates that independent reviewers can reliably apply the classification system. Our reviewers had variable levels of experience, yet they were able to cohesively and accurately use this classification system. This classification system may improve consensus in describing intraoperative laryngeal tissue effects and standardize communications among clinicians and trainees alike. Future prospective studies will be aimed at studying the laser parameters required to achieve each of the classifications discussed in this project.

## Data Availability

All data produced in the present study are available upon reasonable request to the authors.

